# Effect of Varespladib in Patients with Neurotoxic Krait Envenoming: A Subgroup Analysis of the BRAVIO Trial

**DOI:** 10.64898/2026.11.24.26368895

**Authors:** Ashish Bhalla, Chitta R. Mohanty, Badal K. Sahu, Stephen P. Samuel, Suresh Selvam, Manu Ayyan S, Chanaveerappa Bammigatti, Anil K. Goel, Medhavi Gautam, Madhu Kumar R, Surendra Kumar, Harish Kumar, Norman L. Beatty, Charles J. Gerardo, Suneil Agrawal, Eileen Shu, Travis Deuson, Michael A. Gertz, Peter D. Akpunonu, Farshad M. Shirazi, Sarah Watkins, Anne-Michelle Ruha, Shawn M. Varney, Justin Arnold, Samuel J. Francis, Adiel Aizenberg, Andrew Micciche, Rylee S. Bledsoe, Ginger Boatright, Suraj C. Oomman, Janet T. Wittes, Rebecca W. Carter, Matthew R. Lewin, Timothy F. Platts-Mills

## Abstract

**Background:** Envenoming by krait (Genus: *Bungarus*) that are widespread through Asia results in over 10,000 deaths annually in India alone. Krait venoms contain presynaptic secretory phospholipase A2 (sPLA2) neurotoxins that target motor nerve terminals and impair acetylcholine release. The resulting flaccid paralysis may progress rapidly leading to airway compromise, respiratory failure and death. Varespladib, an investigational small-molecule inhibitor of sPLA2, has demonstrated rescue efficacy against lethal krait envenoming in animal models.

**Methods:** We report a subgroup analysis of patients with krait envenoming enrolled in BRAVIO, a multicentre, randomized, double-blind, placebo-controlled trial conducted in India and the U.S. Patients received intravenous followed by oral varespladib or placebo, in addition to standard-of-care antivenom. The trial is registered with ClinicalTrials.gov (NCT05717062) and Clinical Trials Registry-India (CTRI/2023/10/058782).

**Findings:** The efficacy analysis included 45 (24 varespladib, 21 placebo) of the 46 randomized patients. The prespecified primary endpoint was time to recovery of 5-second head lift. The mean time was 27.6 hours for varespladib vs 36.2 hours for placebo, difference −8.6 hours, 95% CI −38.3 to 21.1. Illness severity over the first week, measured using area under the curve for the Snakebite Severity Score through Day 7, was lower in the varespladib group (296 vs 464; difference −168, 95% CI −279 to −57.6). Varespladib patients showed a greater early reduction in Snakebite Severity Score over the first six hours than placebo (difference 1.5, 95% CI 0.5 to 2.5). Among intubated, mechanically ventilated patients (n=25), median duration of intubation was 14.9 hours (IQR 11.5 to 23.3) in the varespladib group versus 34.0 hours (IQR 28.0 to 45.0) in the placebo group. Treatment-emergent adverse events occurred in seven varespladib-treated and six placebo-treated patients (29% each for both groups).

**Interpretation:** The consistent findings of earlier neuromuscular improvement, lower illness severity, and shorter duration of intubation suggest that adjunctive varespladib accelerates recovery from severe neurotoxic krait envenoming.

**Funding:** This work was funded by Ophirex, Inc., the U.S. Defense Health Agency (Award no. W81XWH22C0030), and the FDA’s Office of Orphan Products Development (R01 FD007839). Drug manufacturing was funded in part Wellcome Trust (Contract 222964/Z/21/Z). None of the Defense Health Agency, the FDA’s Office of Orphan Products Development or Wellcome Trust had a role in the writing of the manuscript or the decision to submit it for publication.

**Evidence before this study:** We searched PubMed on October 31^st^ 2026, without date restrictions, using the terms (“varespladib” OR “phospholipase A2 inhibitor”) AND (“snakebite” OR “envenoming” OR “krait” OR “*Bungarus*”) to identify clinical and translational studies of secretory phospholipase A2 inhibitors for snakebite envenoming. We also reviewed relevant preclinical studies, trial protocols, and clinical trial reports.

Krait envenoming can cause severe presynaptic neurotoxicity, resulting in neuromuscular paralysis with resultant airway obstruction, respiratory failure and death. Once established, presynaptic neurotoxicity is resistant to antivenom. Preclinical studies have demonstrated that varespladib can prevent or reverse neurotoxicity caused by several medically important krait venoms. The BRAVO and BRAVIO randomised trials evaluated varespladib as an adjunct to standard care, including antivenom. However, clinical evidence regarding neurological recovery and respiratory outcomes specifically in patients with krait envenoming remained limited.

**Added value of this study:** Krait envenoming is nearly unique among neurotoxic snakebites leaving no fang marks and often delayed realization that a bite has even occurred. This post-hoc analysis of the BRAVIO randomised, double-blind, placebo-controlled trial evaluated 45 patients with krait envenoming who were included in the principal efficacy analysis. The average time to initiation of study drug was more than 9 hours after the bite occurred, and all patients received antivenom prior to initiation of study drug. The primary endpoint, time to recovery of 5-second head lift, was numerically shorter with varespladib than placebo, but the difference was not statistically significant (−8.6 hours, 95% CI −38.3 to 21.1). Exploratory analyses showed lower illness severity through Day 7, greater early improvement in Snakebite Severity Score and grip strength, and shorter duration of intubation among the 25 patients requiring mechanical ventilation. The percent of patients experiencing one or more treatment-emergent adverse event proportions was the same in the two treatment groups. These findings provide preliminary human evidence of neurologic and respiratory benefits of adjunctive phospholipase A2 inhibition in krait envenoming.

**Implications of all the available evidence:** Together with preclinical evidence, these findings suggest the potential of varespladib as an adjunct to antivenom and supportive care for neurotoxic krait envenoming which kill upwards of 10,000 people in India, alone, each year. The observed differences in illness severity, neuromuscular function, and duration of intubation are consistent with a potential treatment effect, but the nonsignificant primary endpoint and small sample sizes preclude definitive conclusions regarding efficacy.

Future studies should evaluate treatment timing, duration of mechanical ventilation, and standardized measures of neurological recovery. The potential benefits of earlier administration, including treatment before hospital arrival, remain to be established and require direct evaluation.

## INTRODUCTION

Snakebite envenoming is an important cause of death in tropical and subtropical regions, with neurotoxic envenoming posing particular challenges because bulbar palsies leading to airway obstruction and respiratory paralysis often lead to venom-induced fatalities before patients reach definitive medical care.^1^ In India, kraits (*Bungarus* spp.), a genus of elapid snakes, account for an estimated 11-18% of snakebites, and contribute disproportionately to the estimated 51,000-58,000 snakebite deaths each year.^1–4^ An estimated 48-64% of patients with krait envenoming develop respiratory paralysis requiring mechanical ventilation.^5,6^

The clinical manifestation of krait envenoming results predominantly from presynaptic insults caused by secretory phospholipase A2 (sPLA2) neurotoxins, including β-bungarotoxin, which disrupt synaptic transmission through the depletion of acetylcholine vesicles from motor nerve terminals.^7,8^ Two limitations constrain treatment of krait envenoming. First, it often takes several hours for victims to reach a hospital where antivenom can be administered safely and where advanced airway management and mechanical ventilatory support may be available. Second, unless given early during the course of illness, antivenom has limited ability to prevent the development and progression of presynaptic neurotoxicity.^9,10^

Varespladib is an investigational small-molecule inhibitor of sPLA2 that has not yet received regulatory approval. As well as intravenous administration, it has pharmacologic properties distinct from antivenom that include oral administration via its prodrug, varespladib methyl, and rapid distribution into tissues inaccessible to antivenom.^11^ Experimental animal studies demonstrate that varespladib can prevent and reverse sPLA2-mediated neurotoxicity, including models using venoms from several medically important *Bungarus* species, including the five major Asiatic krait species.^12,13^ Whether varespladib can improve neurologic outcomes in human krait envenoming remains unknown.

The BRAVIO trial was a double-blind, randomised, placebo-controlled trial evaluating varespladib administered in addition to standard-of-care antivenom for snakebite envenoming in diverse settings and geographies of North America and India.^14^ In this subgroup analysis, we evaluated whether adjunctive varespladib improved neurologic recovery and other clinical outcomes in patients with krait envenoming.

## METHODS

### Study Design and Patients

We conducted this multicentre, randomised, double-blind, placebo-controlled trial at eight sites in India and ten in the U.S. The trial protocol was approved by Indian and US regulatory authorities and the Institutional Review Boards (IRB) or Ethics Committees (EC) at each site. An independent data and safety monitoring board oversaw and periodically reviewed safety data from the trial. The trial was registered at ClinicalTrials.gov, NCT05717062, and Clinical Trials Registry-India, CTRI/2023/10/058782.

Elapid envenoming, a subgroup prespecified in the statistical analysis plan, included patients with krait envenoming in India and eastern coral snake envenoming in the United States. The trial enrolled only one patient with eastern coral snake envenoming. Because of differences in the severity of neurotoxicity and standard of care including antivenom treatment, the present analysis was restricted to patients with krait envenoming to provide a more clinically homogeneous and interpretable study population.

Patients with krait envenoming were eligible if they were randomised within 10 hours of the bite or symptom onset and had moderate or severe cranial nerve or skeletal muscle weakness. Site investigators diagnosed krait envenoming based on the clinical syndrome, the circumstances of the bite (e.g., season, time of day, and location where the bite occurred or the patient had been sleeping), and, when observed, the reported appearance of the snake. Patients were excluded if they were pregnant or had a history of cerebrovascular accident, acute coronary syndrome, chronic kidney disease, or chronic liver disease. In India, enrollment was restricted to adults aged 18 years and older.

### Randomisation and masking

Patients were randomly assigned 1:1 to varespladib or placebo using block randomisation with concealed allocation, stratified by snake type, elapid was one of the prespecified strata. Patients, treating clinicians, outcome assessors, and the trial management group remained masked to treatment allocation until completion of the final analysis.

An unmasked research pharmacist, who was not otherwise involved in patient care or outcome assessment, prepared the intravenous study treatment. Varespladib sodium or placebo was supplied to the clinical team in a 250 mL bag of normal saline. Because varespladib sodium can produce slight visible coloration, the infusion bag and intravenous tubing were covered with opaque material to maintain masking.

### Procedures

All patients received standard of care, including antivenom without protocol-imposed delay or restriction on timing or amount administered.

Adult patients assigned to varespladib received a continuous intravenous infusion of varespladib sodium at 0.45 mg/kg/hr for 6 hours. At 6 hours, patients who were clinically stable and able to tolerate oral medication were transitioned to varespladib methyl, beginning with a 500 mg loading dose followed by 250 mg twice daily for a total treatment duration of 7 days. Patients who were unable to take oral medication at 6 hours continued intravenous varespladib until oral treatment could be started.

### Outcomes

In the BRAVIO study, the primary outcome for patients with krait envenoming was time from randomisation to recovery of 5-second head lift, a real-world clinical sign not requiring any advanced technology to measure. The same outcome definition and statistical approach were retained for the present krait-only analysis. Recovery was defined as the ability, while supine, to raise the head from a flat surface and maintain it elevated for at least 5 seconds. Patients able to sustain a head lift of 5 seconds at baseline were excluded from the primary analysis because they had already met the recovery criterion. Head lift was assessed within protocol-specified time windows at 3 h, 6 h, 24 h (Day 2), 48 h (Day 3), and Days 7, 14, and 28. Assessment of head lift was performed in intubated patients; for those receiving sedation, sedative medications were withheld for at least 60 minutes before assessment.

The parent trial specified three key secondary outcomes: (1) area under the curve (AUC) for the 6-component Snakebite Severity Score (SSS; local wound, pulmonary, cardiovascular, haematologic, nervous, and renal system subscores) from baseline to Day 7 among the subgroup of patients randomised within 5 hours of the bite; (2) SSS of 0 at Day 28, defined by a score of zero for the 6-component SSS; and (3) Patient-Specific Functional Scale (PSFS) score at Day 3 (48 hours).^15^ As prespecified in the statistical analysis plan, change in SSS from baseline to 6 hours was evaluated using the 5-component SSS, which did not include the local wound subscore.

Safety was assessed using vital signs, physical examination, laboratory tests, electrocardiograms (ECGs), clinician assessments, and patient-reported symptoms. Adverse events were categorised by system organ class and preferred term using the Medical Dictionary for Regulatory Activities. Concomitant medications and therapies, clinical interventions, duration of hospitalisation, and adverse events were also recorded.

### Statistical Analysis

Because the parent trial specified separate primary efficacy endpoints for elapid and viper envenoming, sample-size requirements were determined separately for the two groups. For elapid envenoming, assuming a 24-h between-group difference in time to recovery of 5-second head lift, a standard deviation of 16 h, and a two-sided α of 0° 05, 20 patients (10 per treatment group) were required to provide 90% power using a two-sample t-test.

One patient randomized to varespladib inadvertently received intravenous placebo followed by oral varespladib. The decision to exclude this patient from the primary analytic population was made and documented within 24 hours of enrollment. To assess the influence of this post-randomization exclusion, a sensitivity analysis was performed that included all randomized patients according to their assigned treatment group (intention-to-treat population; n=46). See **Appendix Table 1** for results of this analysis.

Analyses of the primary and key secondary endpoints were performed in patients with krait envenoming following the approach prespecified in the statistical analysis plan. Continuous baseline characteristics are reported as means. Analysis of covariance and logistic regression were used to compare differences in continuous and dichotomous outcomes, respectively. AUC values were calculated using the trapezoidal rule by averaging scores for each pair of adjacent assessment time points, multiplying this average by the elapsed time between the prespecified mapped time points in hours, and summing these values over the specified assessment period.

Analyses were adjusted for the variables specified in the statistical analysis plan that were applicable to the krait envenoming subgroup of patients. For the primary analysis, time to complete head lift recovery was compared between treatment groups using a randomisation test with baseline head lift as a covariate in two categories: 0 seconds and 1 to 4 seconds. The randomisation test was selected because of the likelihood of very unequal variances between the treatment groups. Outcomes based on SSS were adjusted for baseline SSS, whereas outcomes based on function scores were adjusted for baseline function scores. Unadjusted longitudinal SSS and patient-reported function scores are presented graphically with standard error (SE) and 95% confidence intervals (CIs). Tables in the appendix provide both adjusted and unadjusted values.

For all SSS outcomes, missing data were handled using multiple imputation following prespecified methods. Subscores were imputed separately within treatment groups under a missing-at-random assumption using Markov chain Monte Carlo to achieve a monotone pattern, followed by predictive mean matching to generate 150 complete datasets.^16^ For figures of SSS and function, averages at each time point were calculated with missing data excluded. Among intubated patients, duration of intubation was compared between treatment groups using the log-rank test. The 95% confidence interval for the between-group difference in median duration of intubation was estimated using non-parametric bootstrap methods.

All analyses were performed with SAS version 9.4 or R version 4.5.1. (SAS Institute, Cary, North Carolina, USA).

## RESULTS

Seven of the eight sites in India enrolled a total of 46 patients with krait envenoming (**Figure 1**). Of these, 25 were randomly assigned to varespladib and 21 to placebo.

**Figure 1.**
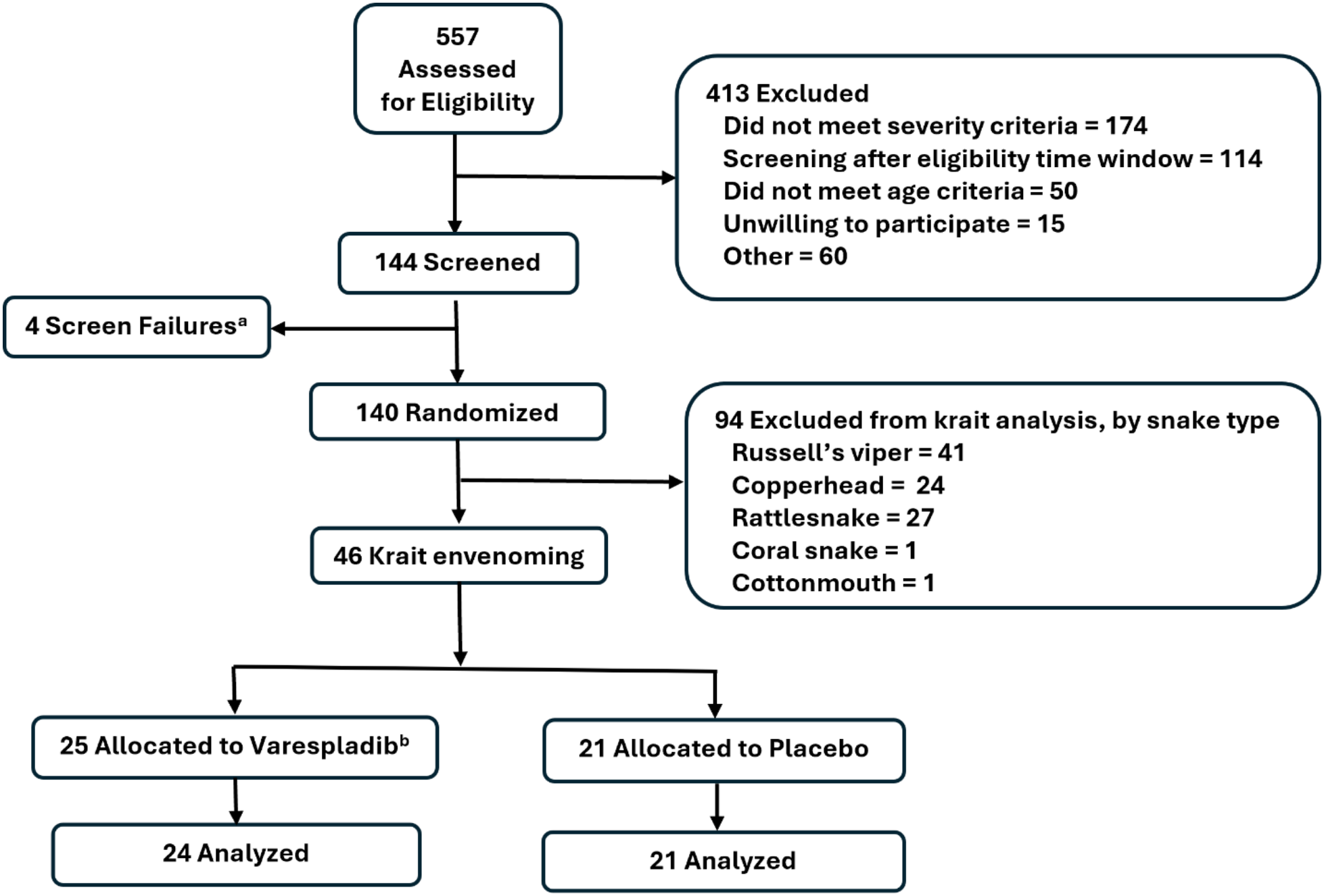
Flow diagram. ^a^ Screen failure reasons: Patient gave verbal consent but then declined to sign consent (n=1); Patient withdrew consent before dosing (n=1); Patient did not meet eligibility criteria (n=2). ^b^ One patient randomised to varespladib received placebo intravenous treatment, then varespladib oral treatment. This patient was excluded from analyses.

Demographic characteristics, baseline illness severity, and baseline neuromuscular function were similar between groups (**Table 1**). All patients received antivenom prior to study drug; the mean elapsed time from antivenom administration preceding study drug treatment was 4.3 hours in the varespladib group and 4.7 hours in the placebo group. Three patients had a baseline head lift duration of 5 seconds; these patients were excluded from the analysis of the primary endpoint but included in other analyses.

**Table 1.** Baseline characteristics.

| **Characteristic** | **Varespladib (n=24)** | **Placebo (n=21)** |
|---|---|---|
| Age, years, mean (SD) | 32.6 (11.0) | 35.1 (12.7) |
| Sex |  |  |
| Male, no. (%) | 15 (62.5) | 15 (71.4) |
| Female, no. (%) | 9 (37.5) | 6 (28.6) |
| SSS<sub>6</sub> at baseline, mean (SD) | 6.6 (2.1) | 6.8 (1.5) |
| PSFS score at baseline, mean (SD) | 2.5 (3.1) | 2.4 (3.4) |
| Grip strength at baseline, kg, mean (SD) | 4.3 (7.4) | 6.6 (9.3) |
| Head lift duration at baseline, no. (%) |  |  |
| 0 seconds | 15 (62.5) | 14 (66.7) |
| 1-4 seconds | 7 (29.2) | 6 (28.6) |
| 5 seconds | 2 (8.3) | 1 (4.8) |
| Timing |  |  |
| Time from bite to antivenom, h, mean (SD) | 5.6 (4.0) | 4.8 (3.6) |
| Time from antivenom to study drug, h, mean (SD) | 4.3 (4.1) | 4.7 (2.8) |
| Total time from bite to study drug, h, mean (SD) | 9.9 (3.9) | 9.5 (3.1) |

SSS<sub>6</sub> = 6-component SSS; PSFS = Patient-Specific Functional Scale

Because the primary endpoint of the BRAVIO trial was not statistically significant, we report nominal p-values. The estimated group difference in time to head lift recovery was −8.6 hours (95% CI −38.3 to 21.1; nominal p-value = 0.29; Table 2). Illness severity over the first 7 days, measured by the 6-component SSS AUC, was lower with varespladib than with placebo (difference −168, 95% CI −279 to −57.6; nominal p-value = 0.003). Improvement in the SSS from baseline to 6 h was greater with varespladib (difference 1.5, 95% CI 0.5 to 2.5; nominal p-value = 0.006), as was the change in grip strength from baseline to 6 hours (difference 6.3 kg, 95% CI 2.5 to 10.1; nominal p-value = 0.001). See Appendix Figure 1 for a forest plot of outcomes reported in **Table 2**. Results for the intention-to-treat population were similar to those of the primary efficacy analysis (Appendix Table 1).

**Table 2.**
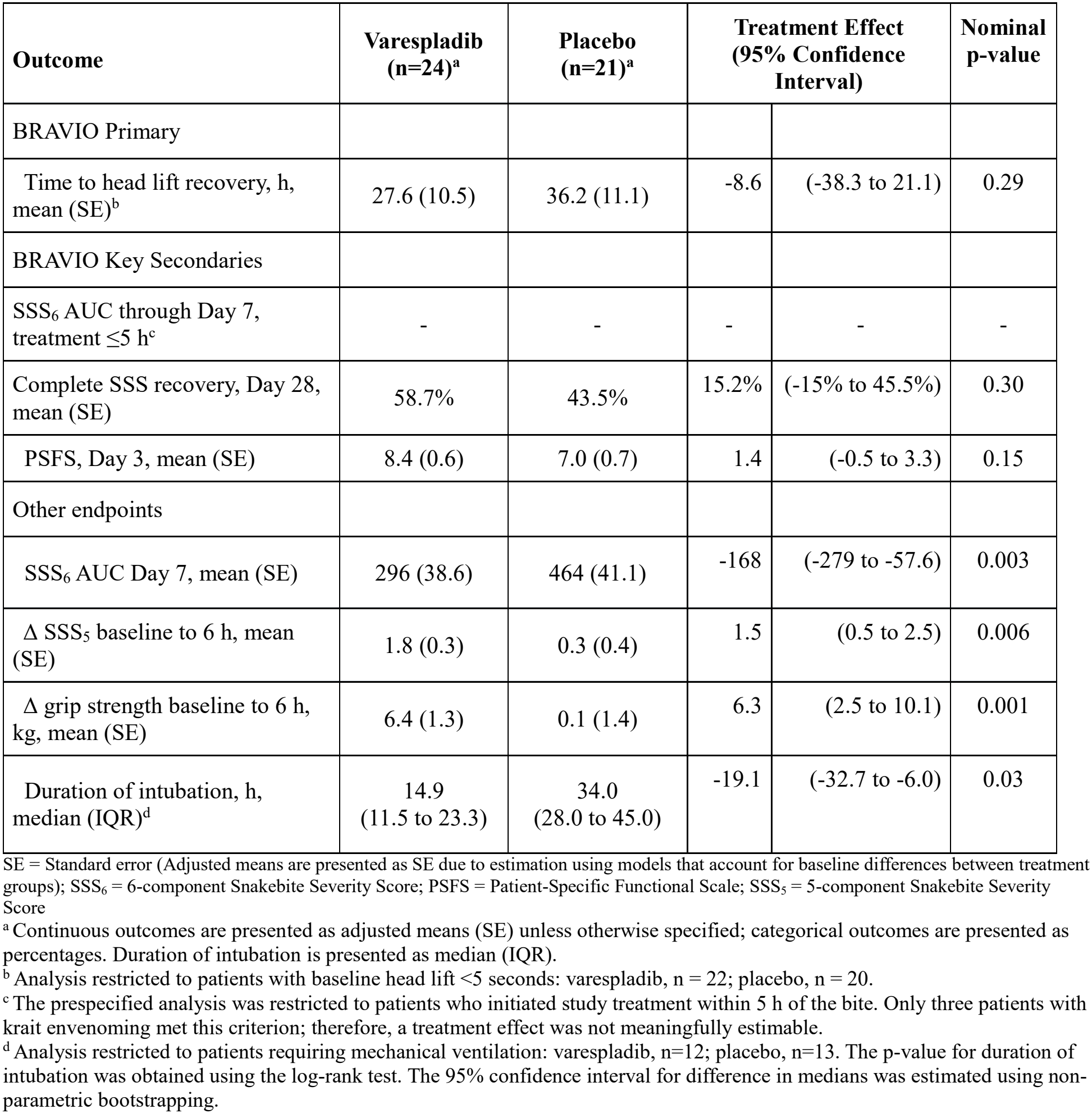
Primary, key secondary, and exploratory endpoints.

Among patients with measurable grip strength at baseline, none of eight who received varespladib had a decline below baseline at 3 or 6 h (or any subsequent time point); five of ten patients assigned to placebo had a decline below baseline in grip strength by 3 or 6 h. Plots of change in grip strength from baseline to 6 h against time from bite to study treatment suggest greater improvements with varespladib than with placebo across a range of treatment times, including among some patients who initiated treatment approximately 18 h after envenoming (**Appendix Figure 2**).

**Figure 2.**
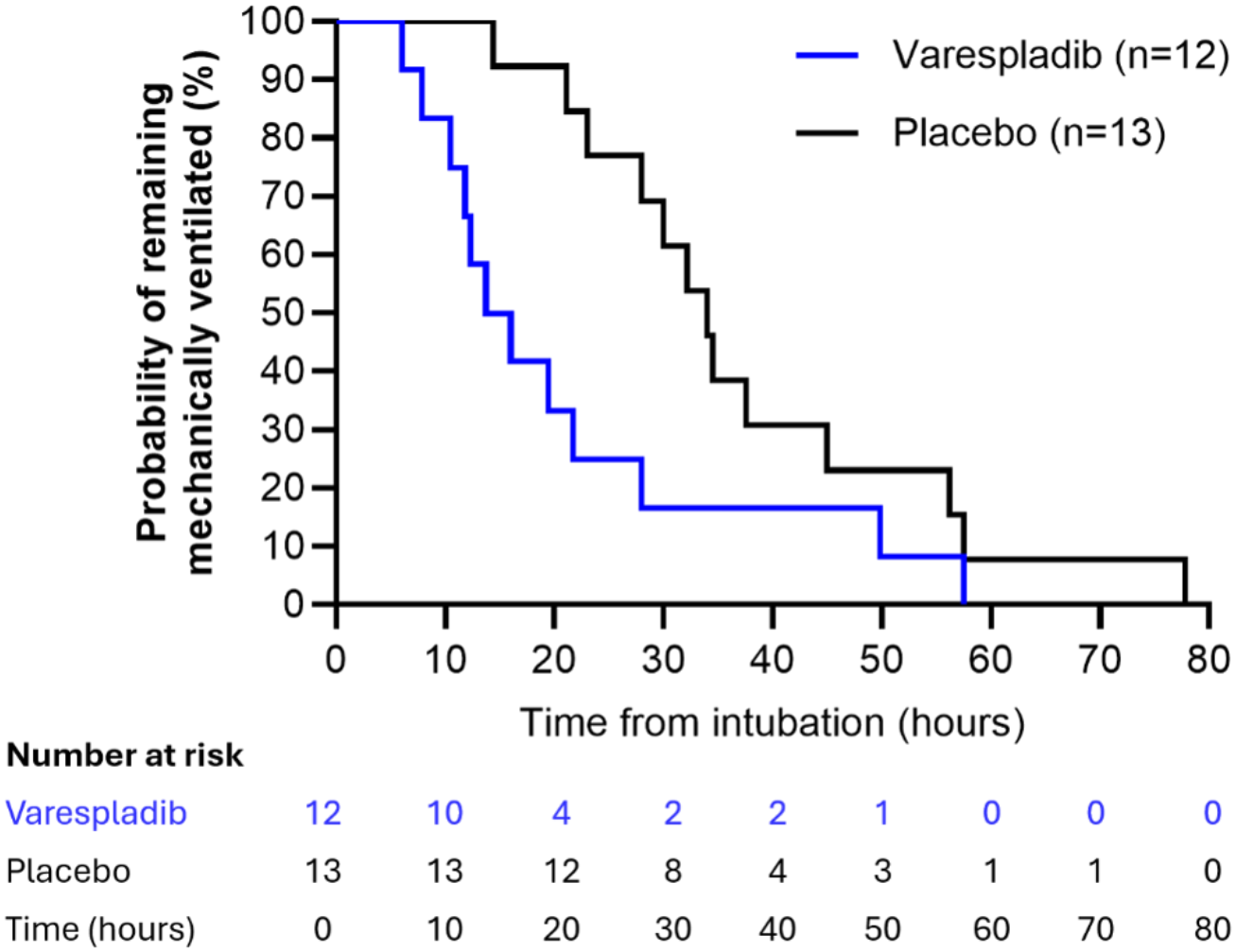
Duration of intubation was significantly reduced among patients requiring intubation and mechanical ventilation. Kaplan–Meier curves are shown for patients assigned to varespladib (n=12) or placebo (n=13). Numbers at risk are shown below the x-axis.

Among the 25 patients requiring mechanical ventilation, duration of intubation was shorter in patients assigned to varespladib (n=12) than in those assigned to placebo (n=13; **Figure 2**). The Kaplan–Meier curves separated early after treatment and remained separated through most of the period of ventilatory support. Median duration of intubation was 14.9 hours with varespladib and 34.0 hours with placebo, a difference of −19.1 hours (95% CI −32.7 to −6.0). The duration of intubation distributions differed between treatment groups (nominal log-rank p-value = 0.03). In the varespladib group, the shortest duration of intubation was 6 hours and 8 of 12 (67%) patients were extubated within 24 hours; in the placebo group, the shortest duration of intubation was 14 hours and 3 of 13 (23%) patients were extubated within 24 hours.

Mean (SD) hospital length of stay was 4.9 (2.5) days in the varespladib group and 5.2 (2.3) days in the placebo group. Mean (SD) intensive care unit (ICU) length of stay was 2.1 (2.0) days and 3.1 (2.3) days, respectively.

Longitudinal trajectories of illness severity and patient-reported function are shown in **Figure 3**. Baseline SSS and PSFS scores were similar between groups. SSS scores separated during the first 6 hours and remained lower with varespladib through 48 hours before converging by Day 7. PSFS scores also improved earlier with varespladib, with separation apparent by 6 hours and persisting through 48 hours; by Day 7, functional scores were similar between groups.

**Figure 3.**
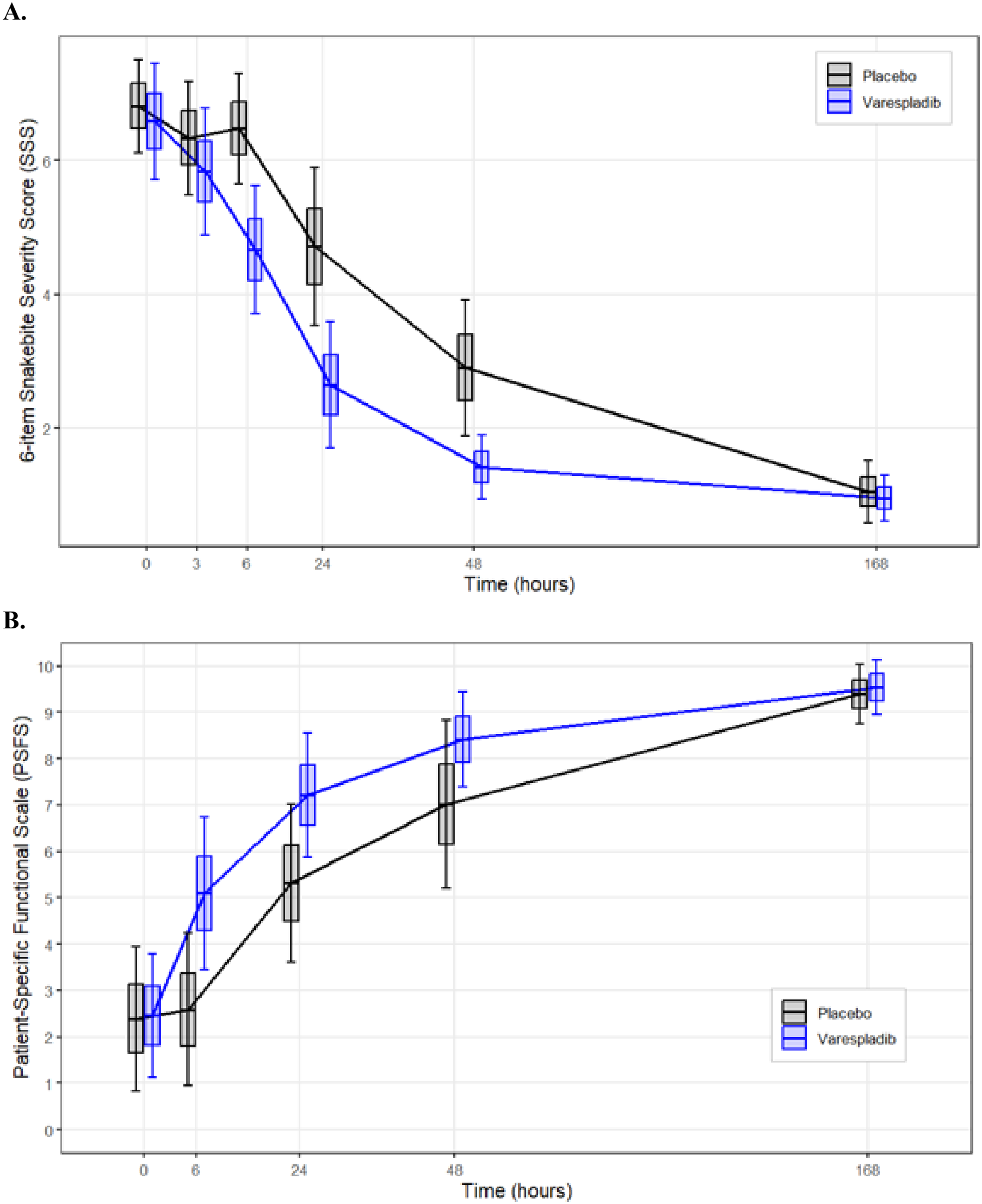
Illness severity and patient-reported function improved over the first 7 days in patients with krait envenoming. (A) Illness severity assessed using the 6-component Snakebite Severity Score (SSS). (B) Patient-reported function assessed using the Patient-Specific Functional Scale (PSFS). Boxes represent the mean ± standard error (SE), with the center line indicating the mean and whiskers representing the 95% confidence interval (CI).

No patient receiving varespladib, either in the krait envenoming subgroup or in the overall trial population died, experienced a serious adverse event, or discontinued treatment. Two serious adverse events occurred in the placebo arm across the entire trial; neither occurred in a patient with krait envenoming. Among patients with krait envenoming, treatment-emergent adverse events occurred in 7 of 24 (29%) patients assigned to varespladib and 6 of 21 (29%) assigned to placebo, comprising 10 and 8 events, respectively. All treatment-emergent adverse events in the varespladib group were mild. The patient randomized to varespladib who inadvertently received intravenous placebo followed by oral varespladib three treatment-emergent adverse events (cough, fever, and nasopharyngitis) on Day 6. All three events were mild in severity. Decreased taste sensation was the most frequently reported adverse event, occurring in five patients assigned to varespladib and three assigned to placebo. Safety results for the overall trial have been published separately;^14^ the appendix table lists all adverse events in the krait envenoming subgroup.

## Discussion

In this subgroup analysis of patients with krait envenoming, varespladib was associated with a consistent pattern of clinical benefit, including earlier recovery of neuromuscular function, substantially shorter duration of intubation, and lower overall illness severity. These findings are consistent with preclinical observations that varespladib can rapidly reverse sPLA2-mediated neurotoxicity in animal models of envenoming in a manner that antivenom does not.^13^ During the first 6 hours, grip strength improved and Snakebite Severity Score decreased in patients receiving varespladib in addition to antivenom whereas these measures were virtually unchanged in patients receiving placebo and antivenom. Collectively, these findings strongly suggest earlier clinical recovery with varespladib, including among patients with severe neurotoxicity requiring mechanical ventilation. The primary endpoint of time to recovery of 5-second head lift was numerically shorter with varespladib but was not statistically significant.

The findings raise an important mechanistic question regarding the reversibility of established presynaptic neurotoxicity. Krait paralysis is predominantly mediated by presynaptically acting secretory phospholipase A2 (sPLA2) neurotoxins such as β-bungarotoxin, which disrupt synaptic transmission by depleting motor nerve terminals of acetylcholine-containing synaptic vesicles and, at least experimentally, cause subsequent nerve-terminal degeneration.^17,18^ The limited ability of antivenom to reverse sPLA_2_-mediated neurotoxicity is thought to reflect the poor accessibility of presynaptic nerve terminals to antivenom antibodies.^10^ Structural studies of snake venom sPLA2 and sPLA2-like toxins show that varespladib occupies the hydrophobic channel, thereby blocking ligand access and constraining structural changes required for toxin activation.^19,20^ Because of its small size, potency, and high permeability, varespladib may have access to tissue compartments that are less accessible to antibody-based antivenoms.^10,21^ Preclinical studies have shown reversal of neurotoxicity within hours in experimental envenoming by *Bungarus* and other presynaptically acting neurotoxic venoms.^11,12,22^ The pattern of clinical recovery observed here is compatible with the hypothesis that a component of established presynaptic dysfunction remains pharmacologically modifiable. The exploratory timing analyses further suggest that varespladib may reverse neuromuscular dysfunction even when treatment is initiated as late as approximately 18 hours after the bite and after flaccid paralysis has developed. Further defining the temporal relationship between toxin exposure, clinical manifestations of envenoming, and reversibility with sPLA2 inhibition will be important for determining the optimal treatment window for varespladib.

The magnitude of the observed treatment effect varied across outcomes, likely reflecting differences in the aspects and timing of neurotoxic recovery captured by each measure. The primary endpoint, recovery of 5-second head lift, was appealing because it is inexpensive, familiar to clinicians, clinically relevant to recovery of axial neuromuscular function and based on low enrollment in BRAVO, there was very concerning uncertainty about the likely number of patients dependent on mechanical ventilation.^14^ However, a 5-second threshold may be relatively insensitive to meaningful improvements in clinical condition. By dichotomising a continuous measure at a single functional threshold, improvement in head-lift duration could occur without satisfying the recovery criterion. Further, because head lift was assessed only at prespecified intervals, recorded recovery reflected the first scheduled assessment at which the criterion was met rather than the exact recovery time. This interval censoring reduced temporal resolution and could have attenuated between-group differences. The wide confidence interval around the primary treatment-effect estimate reflects the relatively small sample size and variability in recovery times.

Krait envenoming classically produces a descending flaccid paralysis, with ocular and other cranial nerve-innervated muscles affected early and weakness then progressing to axial and respiratory muscles and, in severe cases, the limbs.^23,24^ Recovery generally occurs in the reverse order. This pattern may help explain why changes in grip strength were apparent earlier than recovery of 5-second head lift. In contrast to these direct assessments of neuromuscular function, illness severity, patient-reported function, and duration of ventilatory support capture broader dimensions of clinical recovery. These findings reinforce the need to refine and validate clinically meaningful, standardised neurologic and respiratory outcome measures and to select primary endpoints carefully in trials of neurotoxic snakebite.^25–27^

These findings have two potential clinical implications. The most immediate clinical implication is the potential to shorten ventilatory support in patients with established neurotoxic paralysis. If confirmed, this could reduce exposure to ventilator-associated complications and intensive care resource use.^28,29^ A potentially more consequential application is earlier prehospital oral administration of varespladib methyl before progression to respiratory failure, prior to transport or transfer to a hospital capable of advanced airway management.^21,30^ This approach was not evaluated in the present trial; however, the observed effects despite relatively late treatment provide a rationale for prospective evaluation of earlier sPLA2 inhibition. Early treatment could potentially limit progression to severe neurotoxicity and respiratory failure, an important possibility in regions where delays in reaching advanced medical care contribute substantially to snakebite mortality.^1,2^

This study has several limitations. First, although patients with elapid envenoming were a prespecified subgroup, restriction of the present analysis to patients with krait envenoming was post hoc; treatment-effect estimates and p-values should therefore be regarded as exploratory. Second, the sample size was modest, and multiple secondary and exploratory outcomes were examined, increasing the possibility of chance findings, but as a consistent group are highly suggestive of efficacy. Third, krait are nocturnal and identification of krait envenoming was based on its highly stereotypical clinical presentation and epidemiologic characteristics (e.g., season, time of day, patient sleeping location, clinical differences from cobra envenoming that causes local tissue damage) rather than immunologic detection of venom in patient blood which may not be feasible as the toxins are highly neurotrophic. Fourth, study treatment was initiated several hours after the bite and after antivenom treatment. Although rapid initiation of study treatment was prioritised during trial planning and conduct, treatment required patient identification, eligibility assessment, informed consent, randomisation, and study-drug preparation by an unblinded pharmacist, increasing the time between presentation and study-drug administration. The trial therefore does not characterise the effect of early varespladib or varespladib in the absence of antivenom. Finally, decisions regarding intubation, ventilatory management, sedation, and extubation were made as part of clinical care and might have varied between sites. Larger prospectively designed studies will be needed to confirm the observed treatment effects and to define the relationship between treatment timing, neurologic severity, and recovery.

There appeared to be meaningful improvements in ventilator times, and both physician and patient-reported assessments of improvement during the first seven days following severe krait-bite envenoming. In patients with neurotoxic krait envenoming treated with antivenom, the primary endpoint did not show clear evidence that adjunctive varespladib shortened time to recovery of 5-second head lift. However, the evolving consensus measurements and concordant findings of earlier neuromuscular improvement, lower illness severity, and shorter duration of intubation suggest otherwise and support further evaluation of venom sPLA2 inhibition in neurotoxic krait envenoming.^25–27^ Future studies should assess whether earlier sPLA2 inhibition improves recovery from severe neurotoxicity and should incorporate clinically meaningful respiratory, neurologic, and patient-reported outcomes.

## Data sharing

De-identified data for the BRAVIO trial, including the analysis code, and supporting documentation necessary to interpret the data, including data dictionaries and metadata are available via Zenodo (https://doi.org/10.5281/zenodo.19075404).

Any additional data will be made available for related research and analysis upon reasonable written request to the corresponding author within 12 months of the date of publication and subject to appropriate ethics approvals.

## Acknowledgements

We thank the patients for their willingness to participate in the study, those within our organization and at study sites for the dedication required to successfully perform this study, and the DSMB members for their oversight of the study.

## Declaration of generative AI and AI-assisted technologies

The authors used ChatGPT (GPT-5.3 model) for language editing of selected portions of the manuscript. The authors reviewed and edited all content prior to finalizing the submission and take full responsibility for the content of the published article.

## Author Contributions

AB, CJG, SPS, RWC, MRL and TPM were responsible for the original study concept and design for the BRAVO and BRAVIO trials. AB, CRM, BKS, SS, MAS, CB, AKG, MG, MKR, SK, HK, NLB, CJG, SA, ES, TD, MAG, PDA, FMS, SW, AMR, SMV, JA, SJF, AA, AM, RSB screened and enrolled patients into the trials and participated in data collection. JTW and TPM reviewed the statistical analysis. AB, MRL, and TPM had full access to all data in the study and take responsibility for the integrity of the data and the accuracy of the data analysis and serve as guarantors for the manuscript. All authors read and approved the final manuscript.

