## Supplemental Data for "Effect of Varespladib in Patients with Neurotoxic Krait Envenoming: A Subgroup Analysis of the BRAVIO Trial"

### Supplementary Appendix

**Appendix Table 1.** Primary, key secondary, and exploratory outcomes in the intention-to-treat population (N=46).

| Outcome | Varespladib (n=25)[a] | Placebo (n=21)[a] | Treatment Effect (95% Confidence Interval) | |
|---|---|---|---|---|
| **BRAVIO Primary** | | | | |
| Time to head lift recovery, h, mean (SE)[b] | 30.0 (10.1) | 39.4 (10.5) | -9.4 | (-38.9 to 20.1) |
| **BRAVIO Key Secondaries** | | | | |
| $SSS_6$ AUC through Day 7, treatment ≤5 h[c] | - | - | - | - |
| Complete SSS recovery, Day 28 | 58.9% | 44.2% | 14.8% | (-14.3% to 43.9%) |
| PSFS, Day 3 | 8.4 (0.6) | 7.1 (0.7) | 1.3 | (-0.5 to 3.2) |
| **Other endpoints** | | | | |
| $SSS_6$ AUC Day 7 | 288 (37.8) | 463 (41.1) | -175 | (-285.0 to -65.6) |
| Δ $SSS_5$ baseline to 6 h | 1.8 (0.3) | 0.3 (0.4) | 1.5 | (0.5 to 2.5) |
| Δ grip strength baseline to 6 h, kg, mean (SE) | 6.1 (1.3) | 0.2 (1.4) | 5.8 | (2.1 to 9.6) |
| Duration of intubation, h, median (IQR)[d] | 14.9 (11.5 to 23.3) | 34.0 (28.0 to 45.0) | -19.1 | (-32.7 to -6.0) |

[a] The intention-to-treat population includes one patient randomized to varespladib who inadvertently received intravenous placebo followed by oral varespladib, initiated 6 hours and 17 minutes after the start of the intravenous placebo infusion. This patient was excluded from the primary efficacy analysis but was included in this sensitivity analysis according to randomized treatment assignment (varespladib group).
[b] Analysis restricted to patients with baseline head lift <5 seconds: varespladib, n = 23; placebo, n = 20.
[c] The prespecified analysis was restricted to patients who initiated study treatment within 5 h of the bite. Only three patients with krait envenoming met this criterion; therefore, a treatment effect was not meaningfully estimable.
[d] Analysis restricted to patients requiring mechanical ventilation: varespladib, n=12; placebo, n=13. A non-parametric bootstrap method was used to estimate the 95% confidence interval for the between-group difference in duration of intubation.

**Appendix Table 2a.** Unadjusted values for SSS and PSFS, varespladib n=24, placebo n=21.

| | $SSS_6$ | | | PSFS | | |
|---|---|---|---|---|---|---|
| | **Varespladib Mean (SE)** | **Placebo Mean (SE)** | **Unadjusted Mean Difference (SE)** | **Varespladib Mean (SE)** | **Placebo Mean (SE)** | **Unadjusted Mean Difference (SE)** |
| Baseline | 6.6 (0.4) | 6.8 (0.3) | -0.2 (0.6) | 2.5 (0.6) | 2.4 (0.7) | 0.1 (1.0) |
| 3 hours | 5.8 (0.5) | 6.3 (0.4) | -0.3 (0.4) | - | - | - |
| 6 hours | 4.7 (0.5) | 6.5 (0.4) | -1.7 (0.5) | 5.1 (0.8) | 2.6 (0.8) | 2.5 (1.1) |
| Day 2 | 2.7 (0.4) | 4.7 (0.6) | -1.9 (0.7) | 7.2 (0.7) | 5.3 (0.8) | 1.9 (1.0) |
| Day 3 | 1.4 (0.2) | 2.9 (0.5) | -1.5 (0.5) | 8.4 (0.5) | 7.0 (0.9) | 1.4 (1.0) |
| Day 7 | 1.0 (0.2) | 1.1 (0.2) | -0.1 (0.3) | 9.5 (0.3) | 9.4 (0.3) | 0.2 (0.4) |
| Day 14 | 0.6 (0.1) | 0.9 (0.2) | -0.3 (0.2) | 9.6 (0.2) | 9.8 (0.2) | -0.1 (0.3) |
| Day 28 | 0.5 (0.1) | 0.7 (0.2) | -0.2 (0.2) | 9.8 (0.2) | 10.0 (0.0) | -0.2 (0.2) |

$SSS_6$ = Snakebite Severity Score, 6-component. PSFS = Patient-Specific Functional Scale.

**Appendix Table 2b.** Adjusted values for SSS and PSFS, varespladib n=24, placebo n=21.

| | $SSS_6$ | | | PSFS | | |
|---|---|---|---|---|---|---|
| | **Varespladib Mean (SE)** | **Placebo Mean (SE)** | **Adjusted Mean Difference (SE)** | **Varespladib Mean (SE)** | **Placebo Mean (SE)** | **Adjusted Mean Difference (SE)** |
| Baseline | 6.6 (0.4) | 6.8 (0.3) | -0.2 (0.6) | 2.5 (0.6) | 2.4 (0.7) | 0.1 (1.0) |
| 3 hours | 5.9 (0.3) | 6.2 (0.3) | -0.3 (0.4) | - | - | - |
| 6 hours | 4.7 (0.3) | 6.4 (0.4) | -1.7 (0.5) | 5.1 (0.5) | 2.6 (0.6) | 2.5 (0.8) |
| Day 2 | 2.7 (0.5) | 4.7 (0.5) | -1.9 (0.7) | 7.2 (0.6) | 5.3 (0.7) | 1.9 (0.9) |
| Day 3 | 1.4 (0.4) | 2.9 (0.4) | -1.5 (0.5) | 8.4 (0.6) | 7.0 (0.7) | 1.4 (0.9) |
| Day 7 | 1.0 (0.2) | 1.1 (0.2) | -0.1 (0.3) | 9.5 (0.3) | 9.4 (0.3) | 0.1 (0.4) |
| Day 14 | 0.6 (0.1) | 0.9 (0.1) | -0.3 (0.2) | 9.6 (0.2) | 9.8 (0.2) | -0.1 (0.3) |
| Day 28 | 0.5 (0.1) | 0.7 (0.1) | -0.2 (0.2) | 9.8 (0.2) | 10.0 (0.2) | -0.2 (0.2) |

$SSS_6$ = Snakebite Severity Score, 6-component. PSFS = Patient-Specific Functional Scale. Baseline values are unadjusted. Post-baseline values are adjusted for baseline score.

**Appendix Table 3.** Treatment-emergent adverse events by treatment group. All adverse events were mild in severity. A total of eight patients spanning both groups reported decreased taste sensation in the first two days. Though unreported in the literature, taste is regulated by bulbar nerves VII, IX and X.

| **Adverse Event** | **Varespladib (n=24)** | **Placebo (n=21)** |
|---|---|---|
| Any adverse event | 7 (29%) | 6 (29%) |
| Decreased taste sensation | 5 | 3 |
| Fever | 2 | 1 |
| Diarrhea | 1 | 0 |
| Dry mouth | 1 | 0 |
| Vertigo | 1 | 0 |
| Upper respiratory infection | 0 | 1 |
| Chickenpox | 0 | 1 |
| Elevated fibrinogen | 0 | 1 |
| Elevated liver enzymes | 0 | 1 |

**Appendix Figure 1.** Varespladib appears to broadly improve treatment effects versus placebo across clinical outcomes in patients with krait envenoming (n=45).

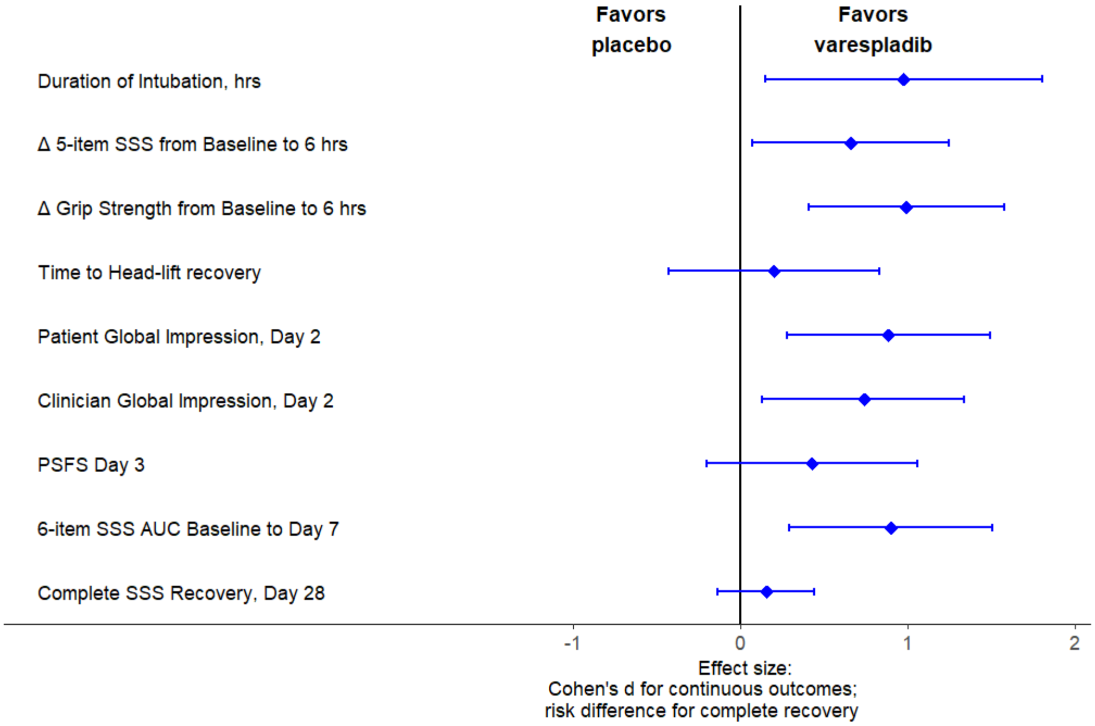


For duration of intubation, the difference between group medians and its 95% confidence interval were estimated using nonparametric bootstrap methods and subsequently standardised for presentation in the forest plot.

**Appendix Figure 2.** Varespladib appears to accelerate return of grip strength from baseline compared to the initiation of study drug treatment among patients with krait-bite envenoming (n=45).

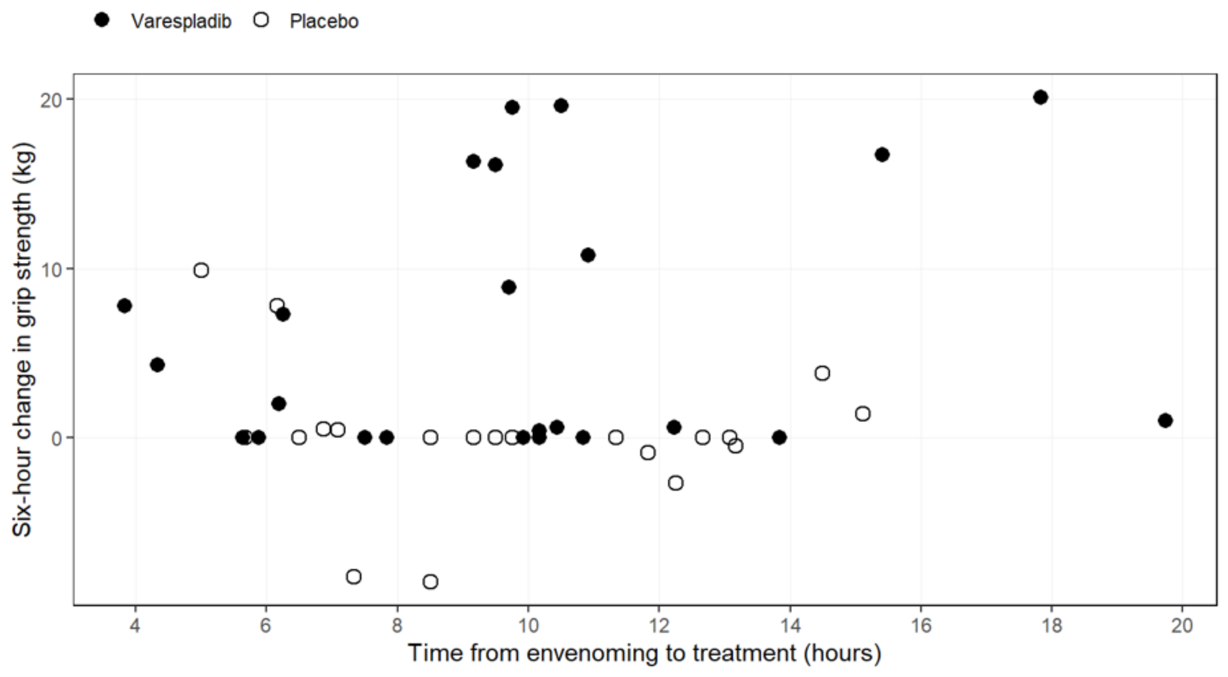
